# Exploring Drug Repurposing for Rare Diseases: Leveraging Biomedical Knowledge Graphs and Access to Scientific Literature

**DOI:** 10.1101/2024.12.31.24319817

**Authors:** Anton Yuryev, Alex Tropsha, Grant Mitchell

**Affiliations:** Elsevier; University of North Carolina; Every Cure

## Abstract

Drug repurposing presents a potential solution for finding new therapies for rare and orphan diseases. The limited number of patients affected by rare diseases, combined with scarce research and the financial burden of clinical trials, creates a significant barrier to developing new drugs. Drug repurposing utilizes the known safety profile and effectiveness of existing medications to fast-track the development of life-saving therapies.

Recently drug repurposing has focused on utilizing biomedical knowledge graphs to uncover hidden connections between diseases and drugs, revealing promising candidates for repurposing. Because most knowledge graphs in biomedical domain are made by text-mining scientific literature we decided to compare the amount of knowledge contained in open access and controlled (subscription only) access literature.

Elsevier and Every Cure make logical partners and allowed the project to use Elsevier’s ability to access both controlled and open access publications and its proprietary Elsevier AI technology to construct the knowledge graph. Notwithstanding the fact that more than 50% of relationships in drug repurposing for rare diseases can be found in open access content, 45% of relationships remain only in controlled access. We argue that this is due to the large number of edges supported by single reference in the entire biomedical knowledge graph and does not reflect an intrinsic difference between open and controlled access.

## Introduction

Drug repurposing, also known as drug repositioning or reprofiling, is the process of identifying new therapeutic uses for existing drugs that were originally developed for a different medical indication. Instead of developing entirely new drugs from scratch, the potential of existing drugs to treat different diseases or conditions is exploited. This approach can significantly reduce the time and cost associated with drug development. Repurposing drugs account for approximately one-third of all drug approvals and almost 25% of overall pharmaceutical sector revenue ^i^.

Examples of successful drug repurposing include:

- finding leprosy and several cancer types as indication for thalidomide that was originally developed as an anti-nausea drug (^ii^)
- repurposing sildenafil (Viagra) initially developed for angina to treat pulmonary hypertension and impotency ^iii^.
- Rituximab (Rituxan): Used for certain cancers, it is now being investigated for rare autoimmune diseases ^iv^.

In the context of rare diseases, drug repurposing holds particular significance ^v, vi^. Rare diseases often have limited treatment options and developing new drugs specifically for these conditions can be challenging due to the small patient populations involved. By repurposing existing drugs, researchers can expedite the identification of potential treatments for rare diseases. This is especially crucial because patients with rare diseases often face delays in accessing effective therapies, and drug repurposing offers a more efficient way to address their medical needs. Additionally, repurposing known drugs may bypass some of the early stages of drug development, such as safety testing, as the safety profile of these drugs is already established. Given that the number of known rare diseases is greater than the number of druggable targets in the human genome, drug repurposing should have a high probability of success, providing right repurposing strategies.

A knowledge graph is a representation of knowledge as a graph structure, where entities (such as drugs, genes, diseases) are nodes, and relationships between them are represented as edges. In the context of drug repurposing, knowledge graphs integrate diverse types of biomedical associations such as protein-protein interactions; drug indications, toxicities, and targets; disease symptoms, clinical parameters, complications, and biological processes; molecular disease biomarkers and other protein-disease associations. Data sources for knowledge graph constructions include scientific literature, clinical trials, public and proprietary databases. Knowledge graphs embedding via machine learning for prediction of new drug-disease connections has become a popular method for drug repurposing ^vii^,^viii^. The embedding is expected to uncover hidden associations between biomedical concepts to find potential new uses for existing drugs.

Biomedical literature is divided into open access and controlled access publications. While historically scientific literature was available by subscription only in controlled access, the open access movement that was started by the scientific community has gained momentum in the beginning of this century. Now open access and controlled access biomedical literature have roughly equal number of articles and the question of relative value of these to corpuses for analytical purposes appears naturally. Understanding of this issue is especially critical for comprehensive efforts in drug repurposing of rare diseases where disease-specific literature is scarce, which makes gathering as much information as possible critical for successful outcome. Elsevier builds its biology knowledge graph from both controlled and open access publications using its proprietary AI NLP technology. Therefore, its graph is a particularly good resource for comparing contributions of two corpuses to biomedical knowledge.

### Definitions

**Observation** (or fact) – relation (causative, physical interaction, or correlation) between two or more biomedical concepts, such as Protein, Biological process, Disease, Cell types etc., that was reported by scientific article. One article can report/reference none, one, or several observations.

**Uncited observation** (uncited relation) – observation supported by single reference in the knowledge graph. Such observations can appear in literature from uncited original research articles or from speculations/hypothesis made by the authors in reviews or in discussions sections of original research articles.

**Statement** – a text snippet describing an observation in a scientific article. Different statements from one or more articles can support a single observation.

**Assertion** (NLP assertion) – statement captured by NLP AI from scientific article supporting observation.

## Methods

### EBKG and its literature sources

The EBKG is made by extracting seventeen types of binary relationships between fifteen types of biomedical and molecular biology concepts. Statistics for each type of node or concept are shown in *Table 1*, statistics for each type of relation or edge is in *Table 2*.

**Table 1:** List of node types and their statistics in current EBKG. Note: The decrease in the number of some entity types in 2023 is due to consolidation and improvements of Elsevier taxonomy.

| Object Type | 2022 |  | 2023 |  |
| --- | --- | --- | --- | --- |
|  | Objects | Aliases | Objects | Aliases |
| Cell Line | 3,412 | 6,115 | 3,416 | 6,128 |
| Cell Type | 864 | 7,331 | 875 | 4,677 |
| Cell Object | 617 | 2,407 | 628 | 2,476 |
| Cell Process | 7,354 | 28,936 | 7,201 | 28,579 |
| Clinical Parameter | 5,404 | 25,950 | 5,669 | 28,292 |
| Disease | 22,929 | 158,795 | 24,264 | 175,601 |
| Compound | 112,438 | 1,020,662 | 113,909 | 1,035,147 |
| Protein/Gene | 139,760 | 733,985 | 144,980 | 744,595 |
| Complex | 979 | 7,377 | 982 | 7,402 |
| Functional Class | 4,587 | 38,646 | 4,970 | 41,349 |
| Organ | 3,839 | 23,458 | 3,972 | 24,607 |
| Tissue | 570 | 3,063 | 582 | 3,188 |
| Virus | 23,597 | 59,056 | 23,763 | 59,912 |
| Pathogen | 607 | 3,106 | 668 | 8,476 |
| External Factor | 73 | 608 | 73 | 608 |
| **Total** | **327,030** | **2,119,495** | **335,991** | **2,171,758** |

**Table 2:**
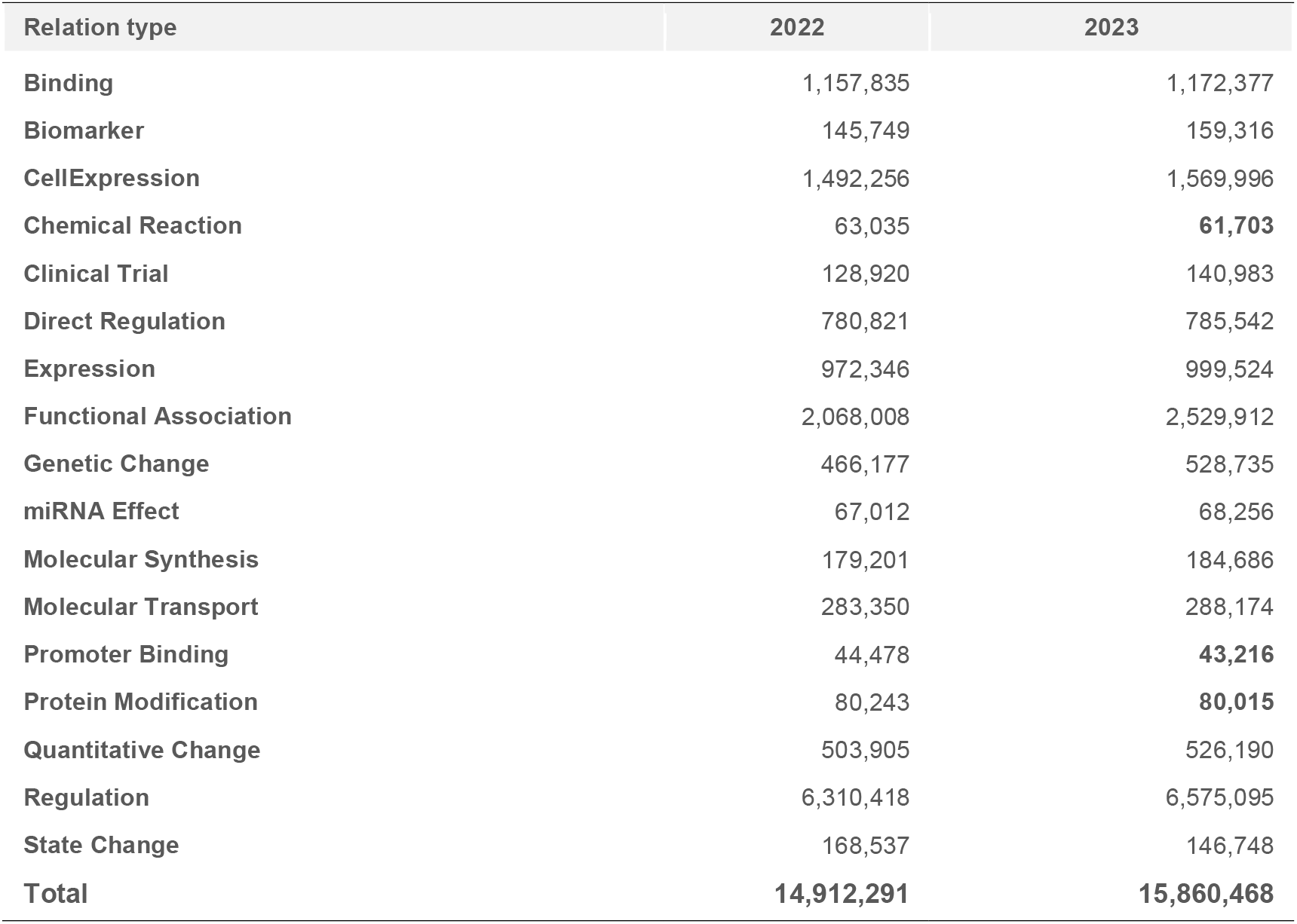
Edge Types and their statistics in current EBKG. The decrease in the number of some relation types in 2023 is due to consolidation and improvements of Elsevier NLP extraction patterns.

| Relation type | 2022 | 2023 |
| --- | --- | --- |
| Binding | 1,157,835 | 1,172,377 |
| Biomarker | 145,749 | 159,316 |
| CellExpression | 1,492,256 | 1,569,996 |
| Chemical Reaction | 63,035 | **61,703** |
| Clinical Trial | 128,920 | 140,983 |
| Direct Regulation | 780,821 | 785,542 |
| Expression | 972,346 | 999,524 |
| Functional Association | 2,068,008 | 2,529,912 |
| Genetic Change | 466,177 | 528,735 |
| miRNA Effect | 67,012 | 68,256 |
| Molecular Synthesis | 179,201 | 184,686 |
| Molecular Transport | 283,350 | 288,174 |
| Promoter Binding | 44,478 | **43,216** |
| Protein Modification | 80,243 | **80,015** |
| Quantitative Change | 503,905 | 526,190 |
| Regulation | 6,310,418 | 6,575,095 |
| State Change | 168,537 | 146,748 |
| **Total** | **14,912,291** | **15,860,468** |

Elsevier uses proprietary AI NLP technology to extract biology knowledge graph from the literature corpus that consists of both open access (OA) and controlled access (CA) sources. The relative amount of each source is shown in Table 3.

**Table 3:** Literature and public database sources for EBKG relevant to drug repurposing. List of journals with full-text articles available to be processed for EBKG can be found in Supplementary file 1.

| Source | OA or CA | Number of documents (millions) | Number of journals |
| --- | --- | --- | --- |
| PubMed abstracts | OA | 34.5 | 14,224 |
| Elsevier full-text articles | CA or OA | 5 | 936 |
| Third party full-text articles | CA or OA | 2.2 | 939 |
| Clinical Trials | OA | 0.43 | N/A |

### AI technology for extracting Elsevier Biology knowledge

Elsevier NLP technology uses named entity recognition and supervised learning for relationship extraction in biomedical literature to create a knowledge graph which empowers researchers with deeper insights from biomedical literature. It employs language models that have been pretrained on large literature corpus consisting of both scientific abstracts and full-text articles. Elsevier NLP accurately identifies genes, proteins, diseases, drugs, and other biomedical concepts, and asserts the semantic associations between them in text. This allows real-time transformation of unstructured text into a structured graph format that can be readily imported into graph or relational database.

Elsevier AI supports entity grounding - extracted concepts are linked to identifiers in external databases, such as NCBI Gene, PubChem, allowing to merge information extracted from the literature with other sources. Every extracted relation is annotated with sentences supporting the relation and identifiers of corresponding articles such as PubMed ID and/or DOI. Additionally, all supporting sentences are labeled by source which allows to differentiate relations extracted from abstracts vs full text. Such comprehensive relation annotation with reference information allows easy assessment of articles availability in OA and CA.

### List of open access publications

List of 8,540,183 PubMed identifiers for OA articles was obtained from Europe PubMed Central website: https://europepmc.org/pub/databases/pmc/DOI/PMID_PMCID_DOI.csv.gz.

### Statistical analysis of EBKG relations

The Elsevier performs disease-centric drug repurposing in two major steps. First, it scores proteins by their importance in disease mechanism. This scoring is done by evaluating the number of articles supporting different types of semantic relations between protein and disease in the knowledge graph (Table 2), by the centrality of a protein in physical interaction subnetwork between proteins linked to disease in scientific literature, and by regulatory potential of each protein relative to disease network. At the second step, algorithm finds drugs modulating activity of ranked proteins and ranks drugs as weighted sum of all scores of its targets obtained at the first step.

Therefore, we selected following two subsets of the knowledge graph as the most relevant for drug repurposing for rare diseases:

- all relations for rare diseases in the knowledge graph (1,025,840 relations for 9,302 rare diseases)
- all drug-target relations in the knowledge graph, including both direct and indirect targets (68,850 direct drug targets; 701,218 indirect drug targets). Indirect drug targets are defined by the literature statements indicating that a protein is regulated by a drug via either unspecified or gene expression mechanisms.

The EBKG contains 1,329,307 relations between chemical substances and their direct protein targets compiled from various public and proprietary Elsevier knowledgebases. Only 4,741 of these relations connect marketed drugs with their targets. We wanted to compare OA and CA contributions into the knowledge graph using relations extracted consistently using one AI technology. We, therefore, excluded 2,390 drugtarget relations that were found exclusively in other knowledgebases and not extracted by Elsevier AI from our analysis.

Rare diseases were found in the knowledge graph as Disease concepts annotated with Orphanet ID. Drugs were selected as Small Molecule concepts under Elsevier ontology categories “drugs” and “plant medicinal product.”

A knowledge graph relation or edge was considered OA if it had at least one supporting OA reference. Otherwise, the relation was considered CA.

## Results

### The contribution of open access facts increases

We first compared the overall contributions of OA (open access) and CA (controlled access) publications to EBKG. The contribution was evaluated by number of NLP assertions supporting relations, the number of publications supporting the assertions, the number of relations and number of journals publishing articles with NLP assertions. We found that contribution of OA and CA publications were the same on the level of knowledge graph relations, while Elsevier NLP asserted 1.5 times more statements in CA corpus compared with OA corpus. The result of the comparison is available in Table 4.

**Table 4:** Contribution of OA and CA publications to EBKG.

| Source | OA | CA | Total |
| --- | --- | --- | --- |
| Number of asserted statements | 29,567,668 | 45,059,137 | 74,626,805 |
| Number of articles | 2,447,164 | 4,986,061 | 7,433,225 |
| Number of relations | 8,456,366 | 7,988,632 | 16,444,998 |
| Number of journals | 13,599 | 4,767 | 18,366 |

We then evaluated the historical trend of OA relations in the portion of the knowledge graph relevant to drug repurposing. The results are shown in Figures 1 and 2. We found that the OA contribution started significantly growing after. 2006. Now more than 50% of all relations necessary for drug repurposing for rare diseases are in OA.

**Figure 1:**
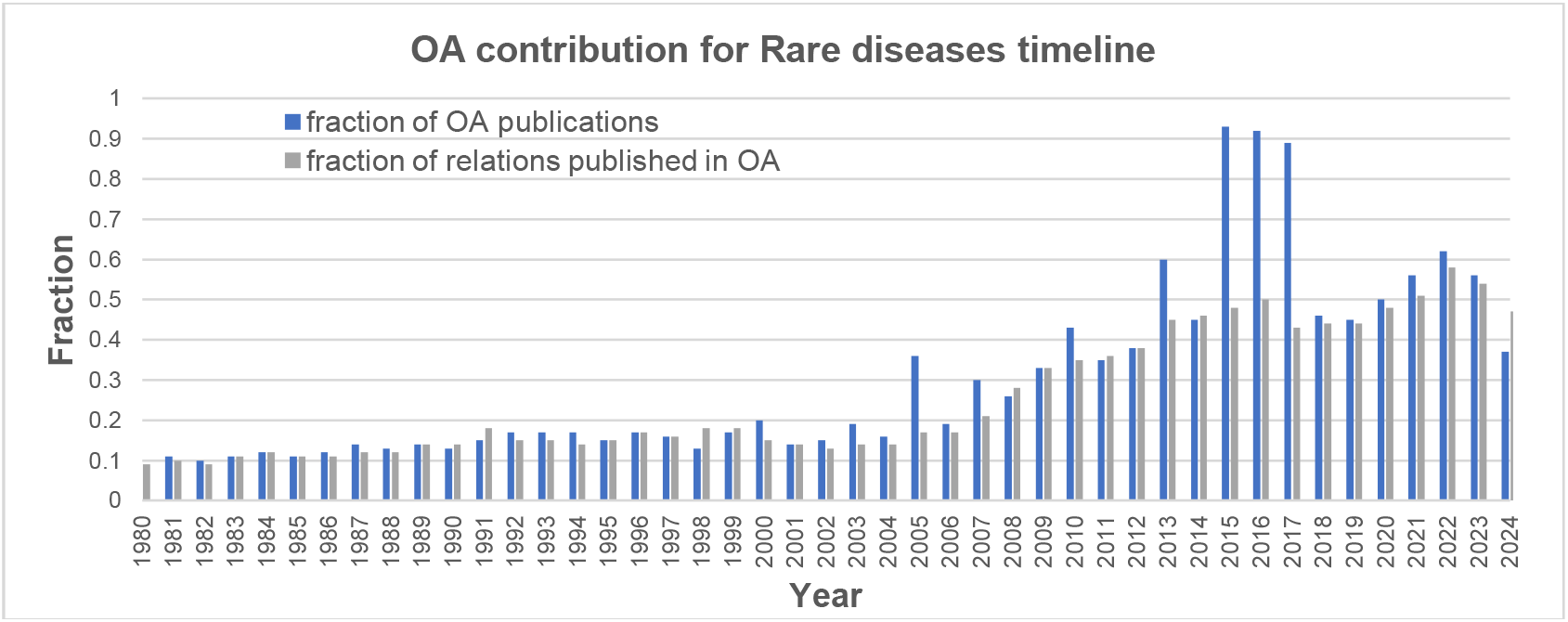
Timeline for OA contribution to rare disease relations since 1980.

**Figure 2:**
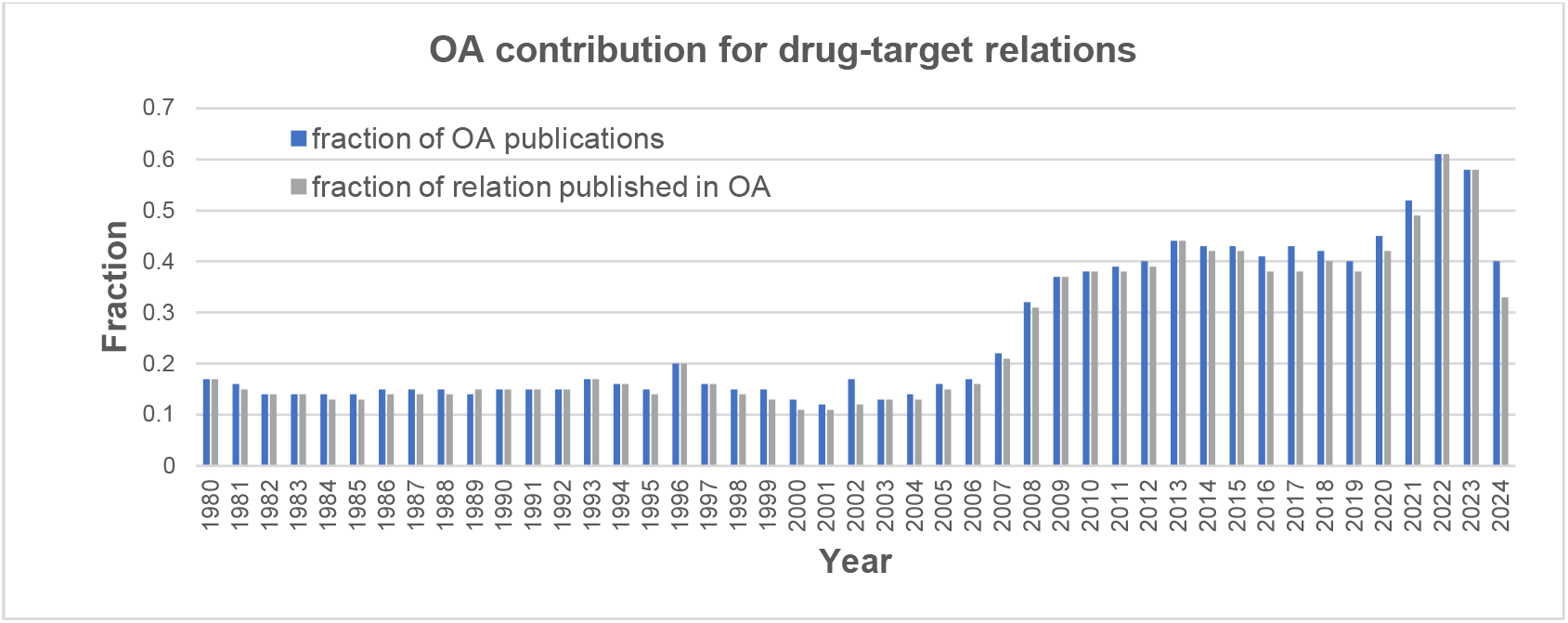
Timeline for OA contribution to drug-target relations since 1980.

The fraction of OA articles is in blue, fraction of OA relations is in red. We did not investigate a reason for large spike in number of OA articles published in 2014-2017 relative to number of published relationships

### The studied diseases have more relations in OA

We then investigated the OA contribution for every rare disease. The scatter plot on Figure 3 shows how disease connectivity or degree correlates with number of OA relations. We found that rare diseases with more than one hundred relations in the knowledge graph tend to have more than 50% of their relations in OA. Interestingly, the similar scatter plot for drug-target relations did not show this bias (Figure 4).

**Figure 3:**
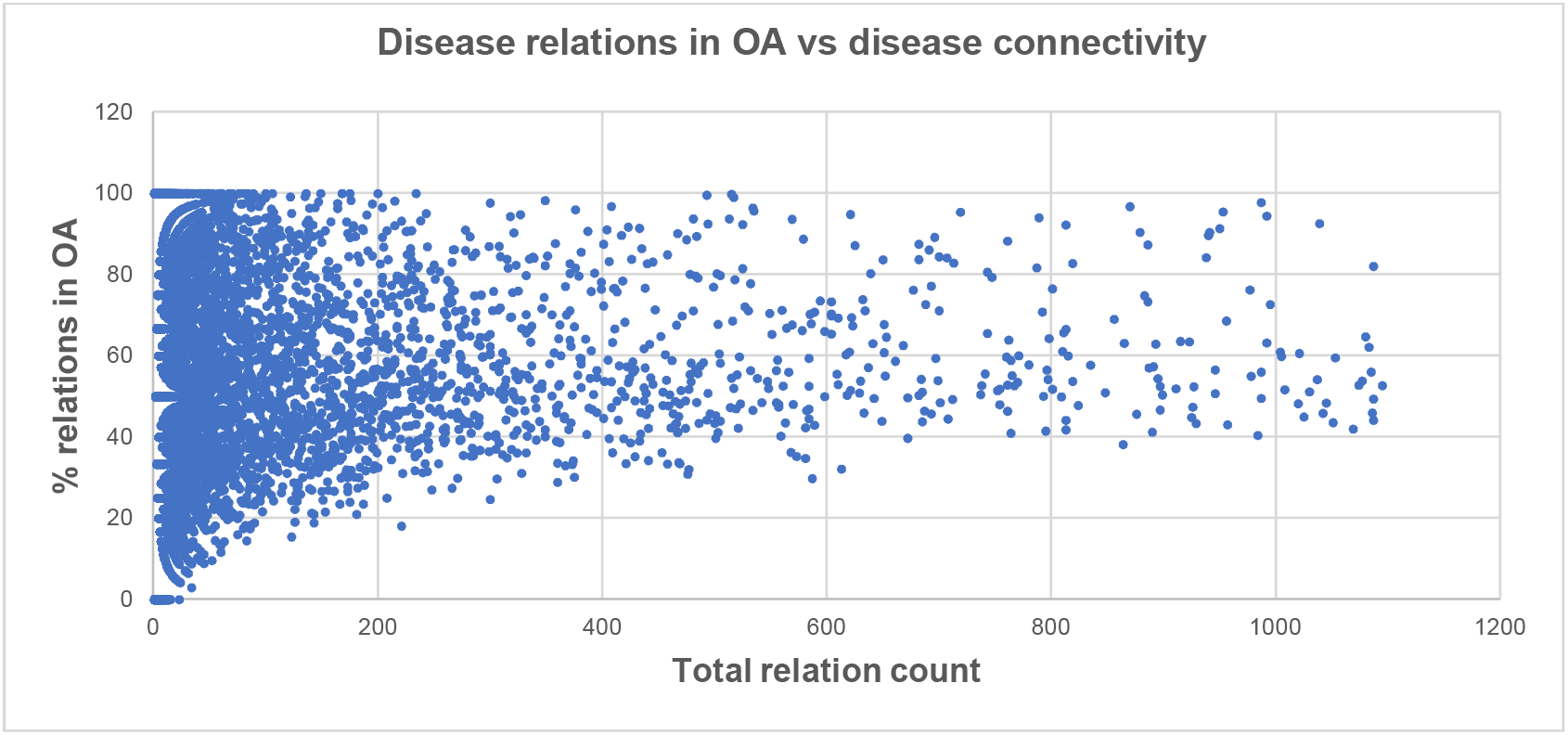
Correlation between percent of OA relations and rare disease research intensity. Each dot on the graph represents one of 9302 rare diseases. Research intensity was estimated as disease connectivity in the knowledge graph.

**Figure 4:**
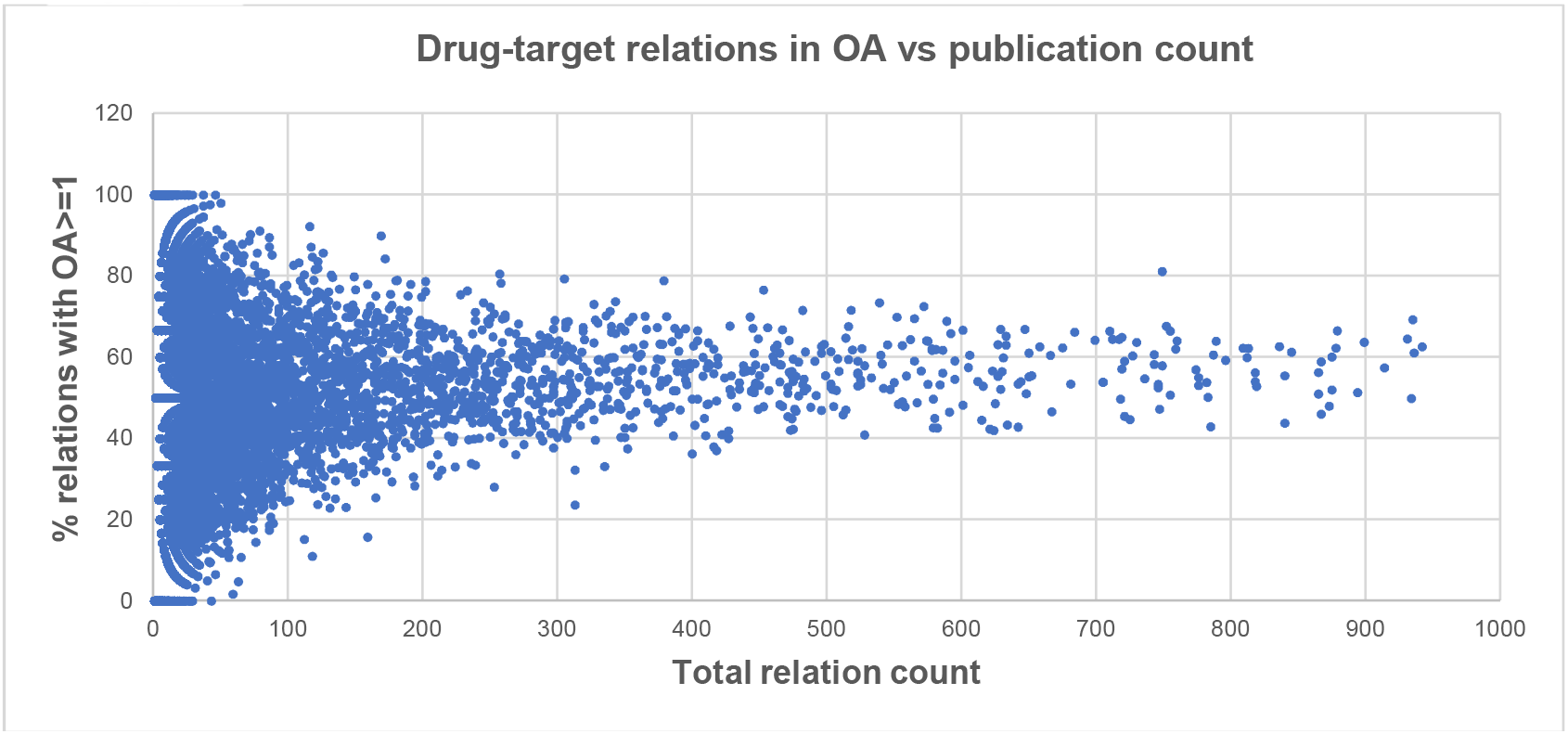
No correlation between OA relations share and drug research intensity in the knowledge graph. Each dot on the graph represents one of 15,744 drugs in EBKG.

### Most relations in both OA and CA are supported by a single reference

We then investigated how many references on average support relations in OA. We first plotted the proportion of OA relations versus the number of supporting references. We found that almost all relations supported by 4 or more references are in OA, while the fraction of OA relations supported by single reference is 40% (Figures 5 and Table 5)

**Table 5:** Fraction of CA relations for rare diseases relations supported by different number of references.

| All relations | Relations supported by 2 or more references | Relations supported by 3 or more references | Relations supported by 4 or more references |
| --- | --- | --- | --- |
| 0.36% | 0.15% | 0.08% | 0.06% |

**Figure 5:**
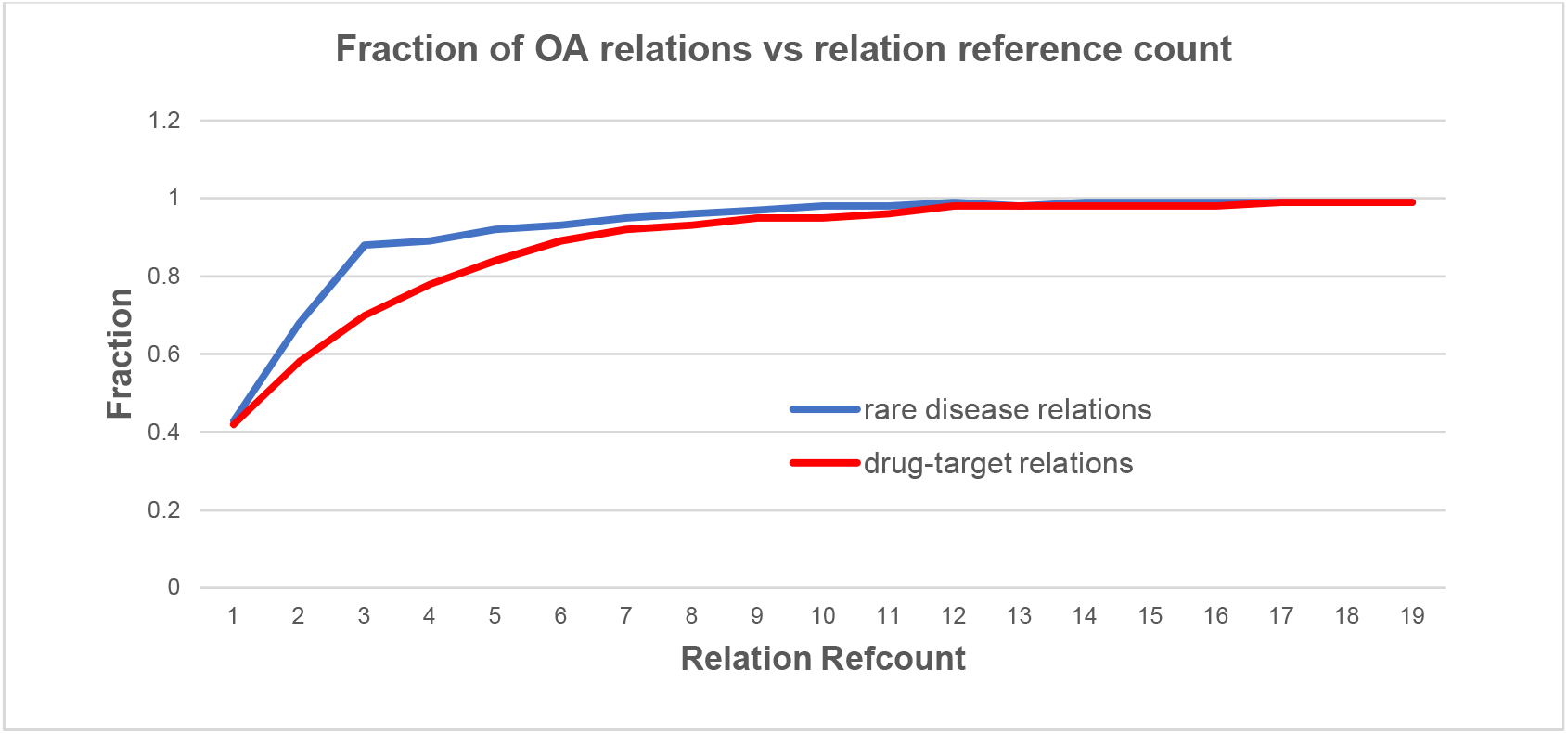
Fraction of OA relations among relations with different number of supporting references (reference count)

### The total number of relations published in CA also increases

The apparent decrease in the CA knowledge contribution in our knowledge graph has prompted us to investigate the growth of the overall CA knowledge over time. We found that the number of facts published in CA articles increases every year and there is no decline int time in the amount of CA only content (Figure 6). The growth of CA only knowledge, however, is slower than the growth of OA knowledge and therefore the apparent contribution of CA content measured as the fraction of CA only relation declines over time.

**Figure 6:**
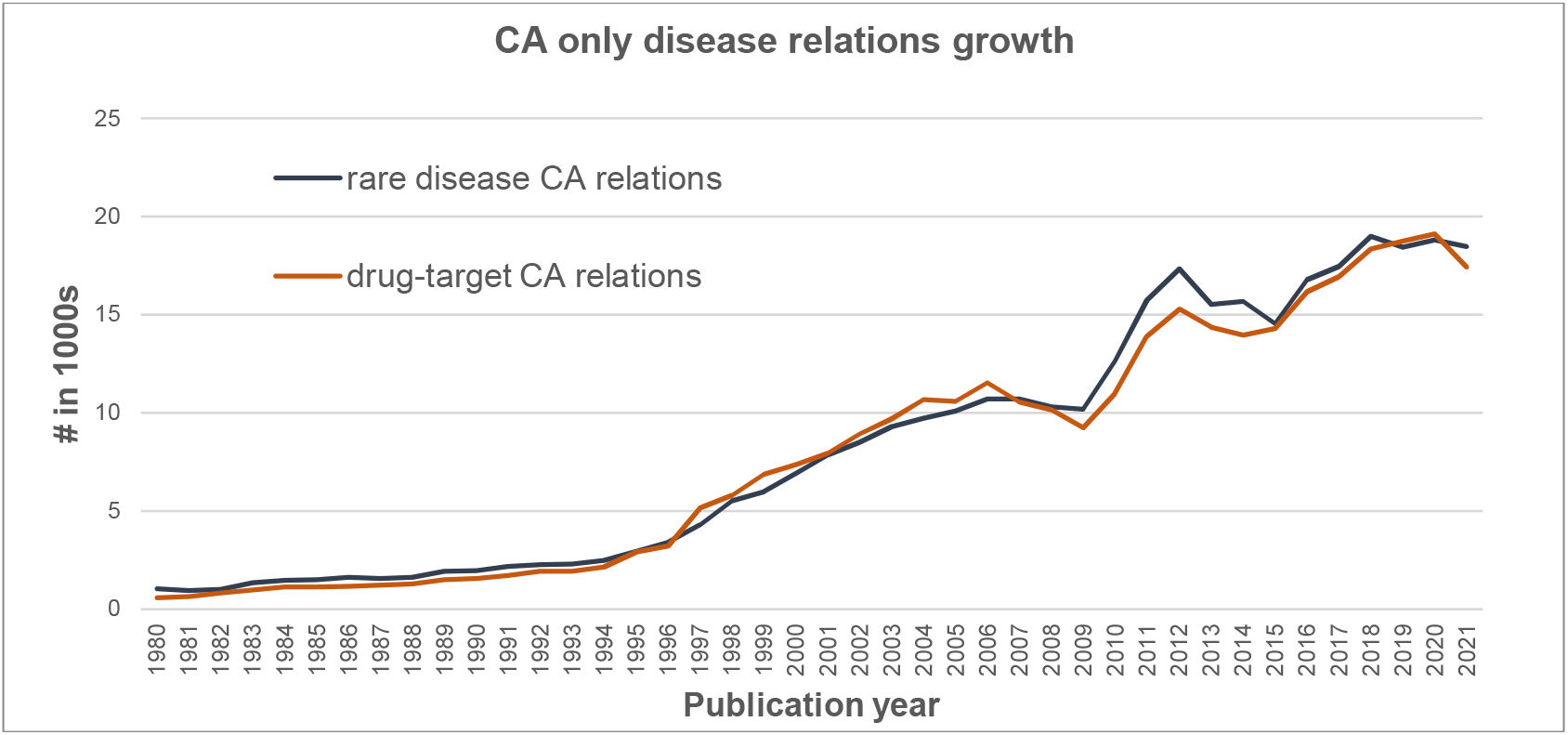
The amount of CA only facts in scientific literature every year.

**Figure 7:**
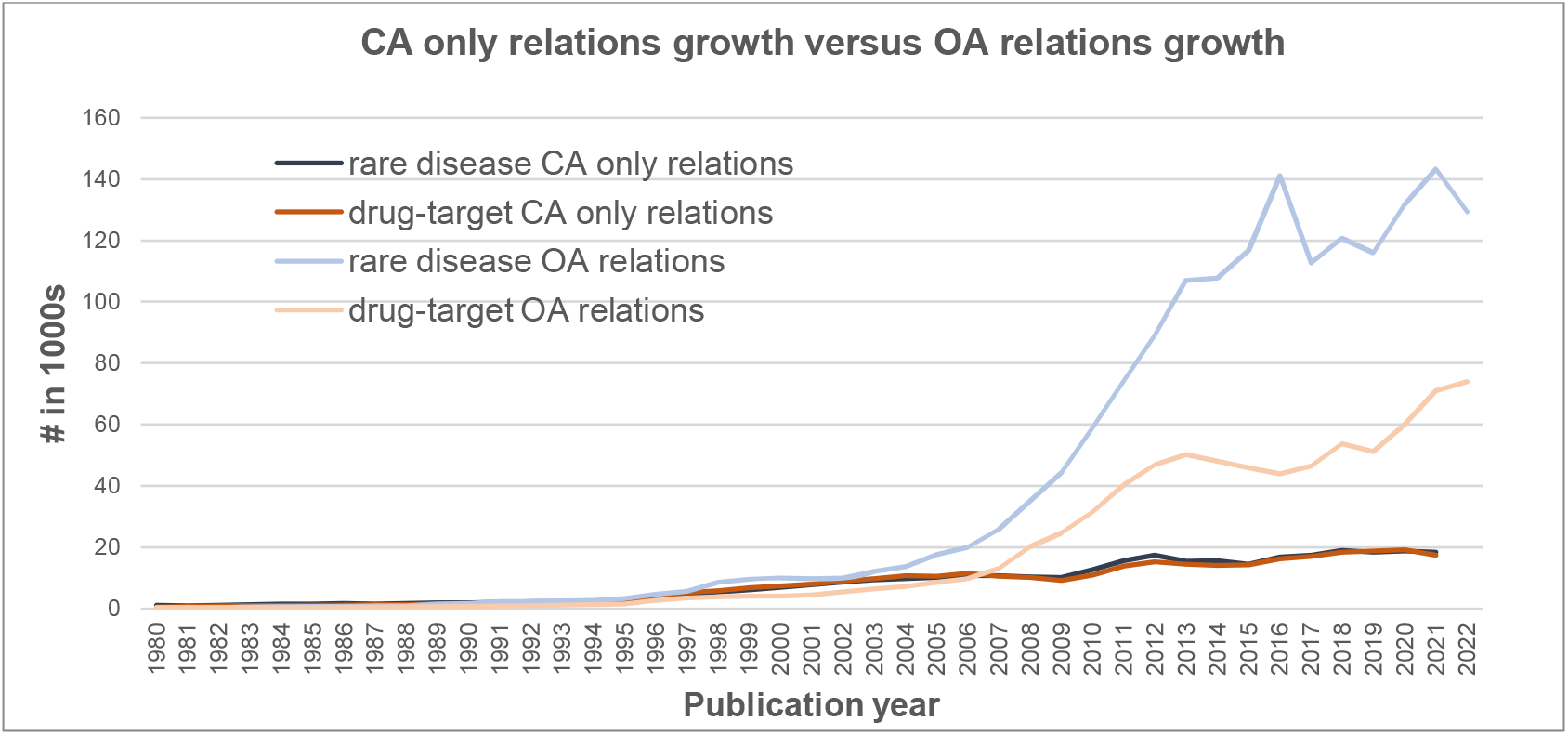
Growth of OA relations is due to both publishing results in OA articles and referencing CA results in OA literature.

### No difference in citation rate between OA and CA

Because CA content is generally less accessible, one possible explanation for the accumulation of CA relations in the graph can be the different citation rate between relations that were first published in OA and relations first published in CA. We compared the average time for the first citation for CA and OA literature sources. The first citation of a relation happens either due to second experimental confirmation of the observation or due to a relation mentioning in support of biological model of disease or drug mechanism in another article. Figure 8 shows that there is essentially no difference in citation rate between OA and CA content. Thus, there is no barrier for discovery of CA relations that could have existed due to more strict accessibility of CA articles. Moreover, graph on Figure 5 shows that the time to the first citation decreases historically for both CA and OA relations, which is expected from the growth of information technologies allowing easier search and retrieval of scientific articles.

**Figure 8:**
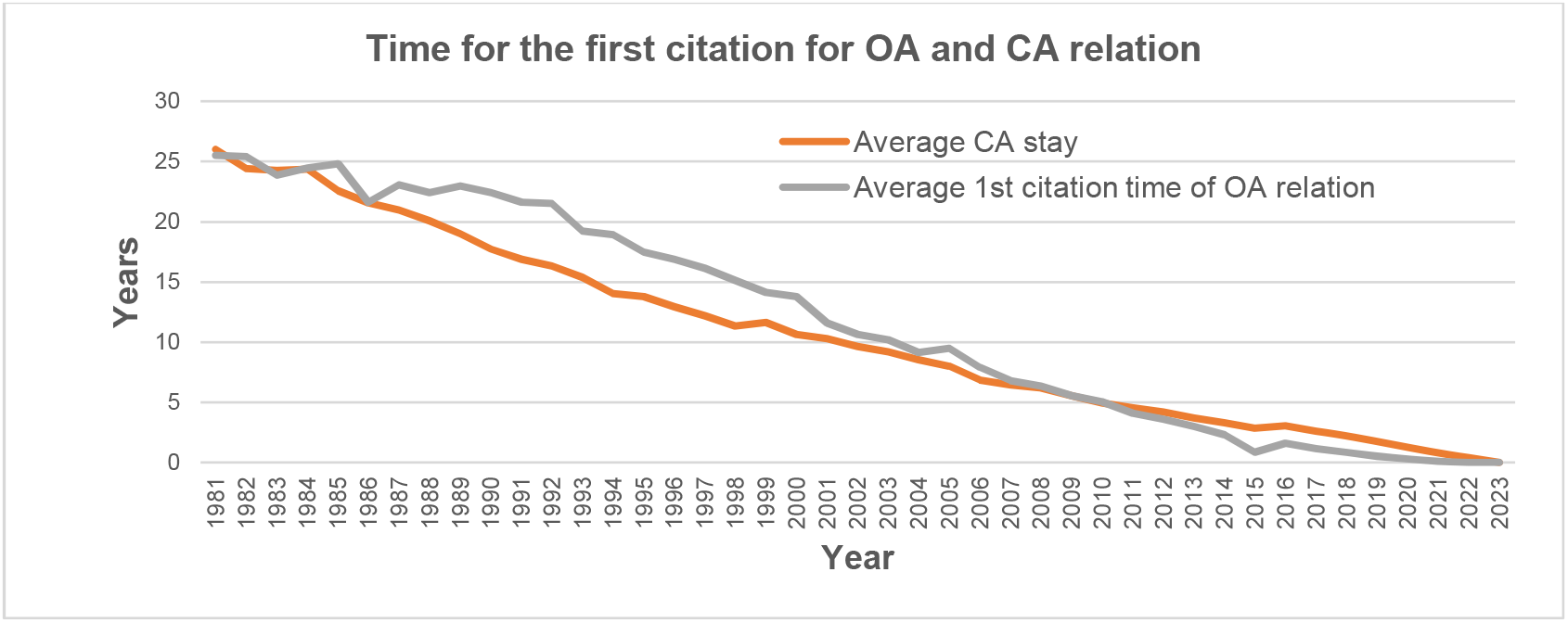
Time to first citation for OA and CA relations published in different years. “Average CA stay” – average amount of time (in years) for a relation published in CA to get first citation in OA.

### Exponential accumulation of single reference relations in scientific literature

The data above shows that despite the advent of OA in 2000 and accelerated growth of OA content since 2006 about half of all relations in EBKG remain in CA. Most of these relations are supported by a single reference. To find whether accumulation of single reference relations is unique property of CA content we have determined the number of rare disease relations supported by single reference published every year. Figure 10 shows an almost exponential accumulation of such uncited relations in time. Thus, rare disease relations supported by single reference are being accumulated in both CA and OA literature corpuses. While relations published recently have generally less chance to be cited, the decrease in time for the first citation shown in Figure 9 suggests that the main reason for accumulation of uncited novel relations in the graph is an absence of the follow-up by scientific community.

**Figure 9:**
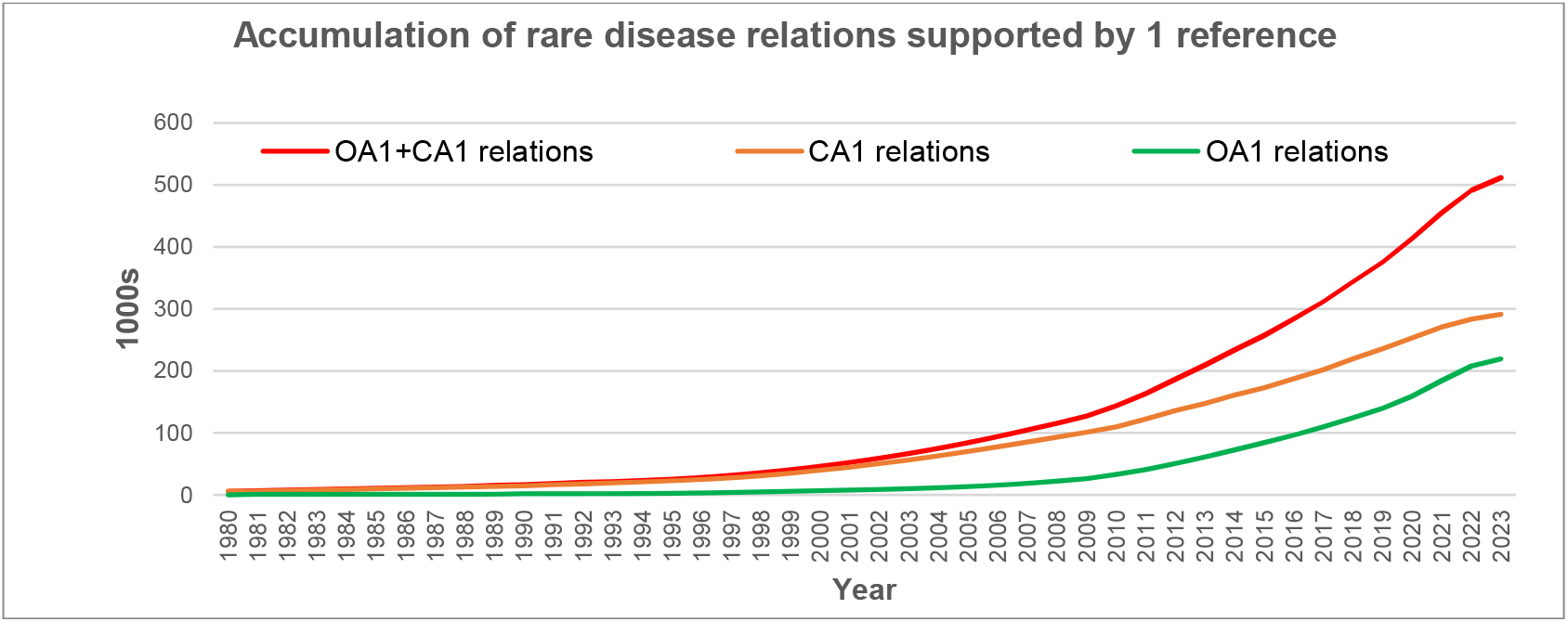
Accumulation of rare disease relations supported by single reference in EBKG. OA1 – relations supported by one reference in open access, CA1 - relations supported by one reference in controlled access.

**Figure 10:**
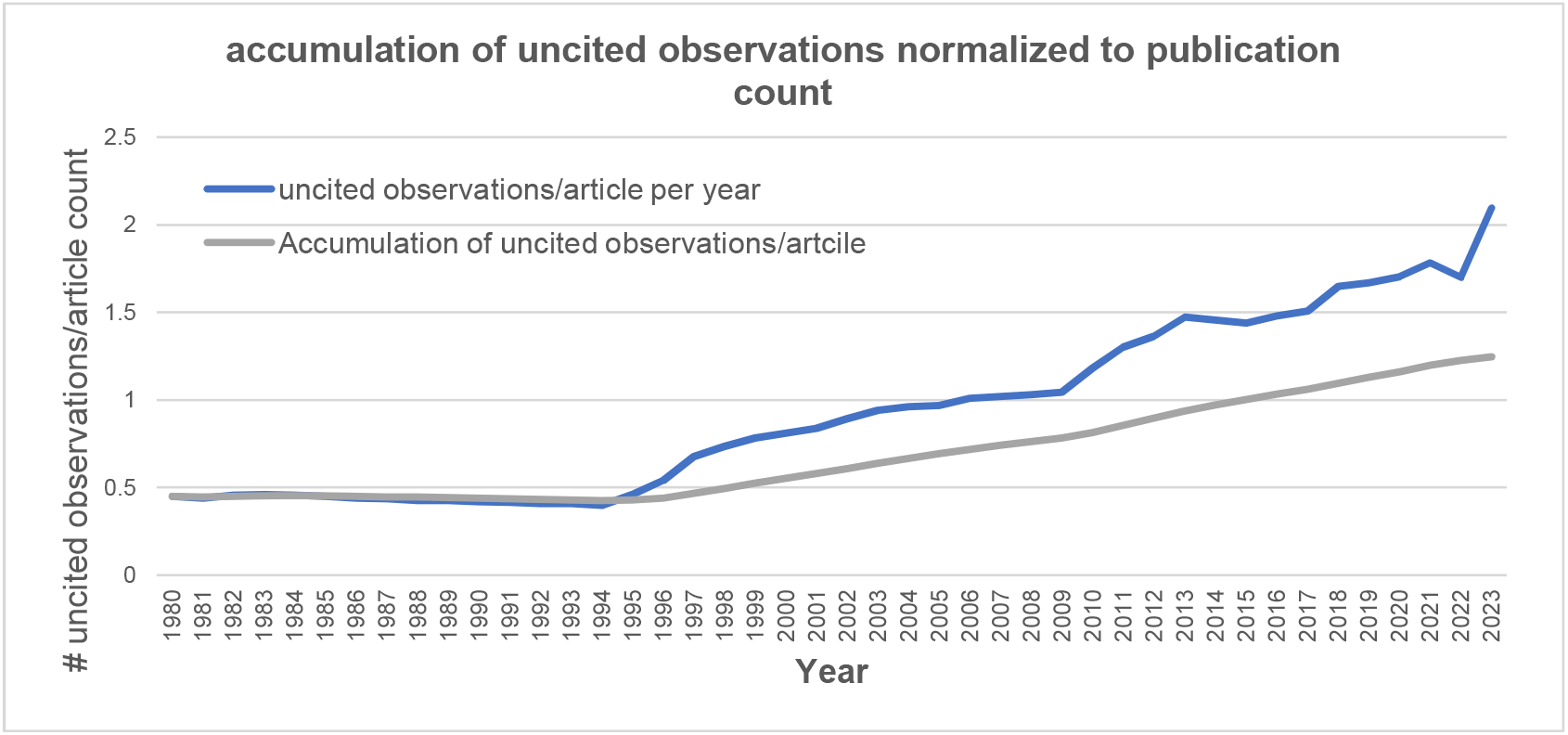
Increased proportion of uncited observations in EBKG. The Accumulation graph shows the ratio of accumulated uncited observations relative to the total number of articles in EBKG every year.

To investigate if the exponential growth of uncited relations is unique to rare diseases or is a general trend in biomedical literature, we plotted the accumulation of single-reference relations in the entire EBKG (Figure 10). We found the same explosion of uncited research in the entire biomedical research.

Besides relations extracted by Elsevier NLP from scientific publications EBKG also contains relations imported from the third-party public databases and high-throughput experiments. To make sure that the observed accumulation of uncited relation is indeed the trend of peer-review literature and not the effect of unverified results from high-throughput experiments we have counted the number of uncited relations produced by different sources in EBKG. Table 6 shows that indeed 85% of uncited relations were extracted from peer-reviewed scientific literature.

**Table 6:** How uncited observations are distributed across knowledge graph sources.

| Relation Source | total # relations | # uncited relations |
| --- | --- | --- |
| Entire graph | 16,635,176 | 9,950,525 |
| Elsevier NLP | 14,459,293 | 8,486,999 |
| Reaxys | 1,329,300 | 1,140,023 |
| ClinVar | 261,265 | 38,445 |
| BioGRID High Throughput | 177,680 | 156,984 |
| BioGRID Low Throughput | 52,074 | 23,256 |
| TargetScan | 19,837 | 16,257 |
| PicTar | 13,959 | 10,290 |
| miRanda | 13,451 | 11,263 |
| DrugBank | 8,243 | 1,779 |

### The explosion of uncited observations in biomedical literature

To understand better the dynamics of expansion of relations supported by single reference we plotted the ratio of the number of uncited relations (observations) reported every year relative to the total number of observations made in the same year. Figure 8 shows that the proportion of uncited relations published by article was dropping steadily until the year 2000. This downward trend stabilized in the beginning of this century and the fraction of uncited relation remained mostly constant. The slight increase of the ratio in the last two years is probably because recently discovered relations did not have enough time to get cited. The observed trend coincides with the growth of the open access movement, which aims to increase the research findings accessibility. Some may argue that open access helped to ensure that important new discoveries are not overlooked by the traditional peer review process ^ix^.

### Higher article retraction rate in CA journals

The lack of citations for a reported relation may stem from low research activity in the field or difficulties in replicating the original findings. Rare disease research, hampered by limited funding and challenges in gathering data from small patient groups, can be particularly susceptible to both low research intensity and poor reproducibility. To explore the impact of reproducibility and potential differences in research quality between openaccess (OA) and closed-access (CA) knowledge, we examined the number of retracted articles in both literature corpuses. Of the 27,000 retracted articles identified in PubMed, only 8,562 (31.7%) were OA publications. This difference might be attributed to variations in peer review rigor, data sharing practices, and data format standards between OA and CA publishing models.

### Most uncited relations in the knowledge graph are from original research articles

There are several reasons for a relation to become uncited. First, Elsevier NLP generates about 10% of false positive relations (Table 7).

**Table 7:** Counts of relations by the number of supporting NLP assertions in knowledge graph.

| Number of relations | 2022 | 2023 | Confidence |
| --- | --- | --- | --- |
| Individual statements | 67.1M | 54.9M |  |
| Edges (relations): |  |  |  |
| •All | 14.9M | 15.9M | > 90% |
| •2 and more refs | 6.0M | 6.4M | > 95% |
| •3 and more refs | 3.8M | 4.1M | > 97% |

One major source of NLP false positives is long complex sentences that list relations between multiple concepts in the form of grammatic conjunction. Such sentences generate multiple NLP assertions some of which can be false positives.

The second reason for relation to be uncited are true positive NLP assertions generated from hypothetical statements that are usually made by authors in the Discussion section of the original research articles or in review articles. Such hypothetical statements may be unproven by subsequent research. However, their refute is never reported due to positive publication bias^x^.

The third reason for the uncited relation may be a true positive observation reported by an original research article that has passed peer review, but its findings have never been followed by the scientific community. Original research articles often contain multiple statements about the main observations that they report. The main findings are usually repeated in Abstract, Results and Discussion sections of an article. Frequently, reported experimental observations are measured by different techniques described in different subsections of the Result section. Therefore, the Result section can also have additional statements supporting reported observation.

To better understand the reasons for explosive growth of uncited research in scientific literature for every article in the knowledge graph we have calculated a ratio between number of NLP assertions made from one article and the number of unique relations asserted from the same article. The ratio bigger than 1 suggests that an article is an original research article reporting one or few relations. The ratio less than 1 suggests that an article is a review containing long complex sentences with relations conjunctions. We then compared the distribution of assertions-to-relations ratio in all articles in the knowledge graph versus distribution of this ratio in the articles reporting one or more uncited relations. Figure 9 shows below the result of comparison. We find that articles reporting uncited relations on average have a higher assertions-to-relations ratio than articles in the entire graph. This suggests that most uncited relations come from the original research articles that are not followed up by the subsequent research.

## Discussion

The OA movement started in the beginning of this century with establishing non-profit scientific publishers such as Public Library of Science and BioMed Central in 2000. The concept of OA was formally introduced in 2001 by the Budapest Open Access Initiative (BOAI) launched by the Open Society Institute. The BOAI sought to establish principles for unrestricted public access to scholarly research. While the definition of OA has evolved with subsequent implementations, it generally represents the intent to enable free and permanent access to published research, often coupled with explicit permissions for reuse and dissemination. Elsevier has two primary open access publishing models: gold OA, where journals publish OA articles funded by author fees, and green OA, where authors self-archive their published articles in open repositories.

The intention of OA founders was to widen societal impact of scientific research by increasing collaboration and information sharing, by reducing barriers to knowledge, enhancing transparency and reproducibility, and faster dissemination of scientific findings^xi^. We found no difference in research citation rate between CA and OA publications. We also found similar fractions of CA and OA get cited at least once indicating that there is no difference in the frequency of citation. The average first citation time is not different between scientific facts first reported in CA or facts reported in OA, indicating that there is no difference in the speed of first citation. We show that while the overall impact of OA on biomedical knowledge steadily increases over time due to the increasing number of OA journals, the knowledge relevant for drug repurposing is equally distributed between OA and CA literature. We attribute this to the exponential growth of biomedical literature in general. This explosive growth leads to the accumulation of relations supported by a single reference in both corpuses. These relations are not being followed up by subsequent research. Indeed, we found that single referenced relations constitute more than 50% of all relations in EBKG (Table 6).

The OA movement came about in part from concerns that the traditional peer review process could limit access and slow down scientific communication. Indeed, we found that the number of novel findings per publication decreased until the year 2000 and then stabilized. The downside of this development was explosive growth of uncited scientific observations and claims that were reported equally in OA and CA literature. While every uncited relation has passed the peer review process the quality of peer reviews may vary with journal. However, the very small fraction of retracted articles compared to the total number of publications reporting novel relation suggests again that the accumulation of uncited relations is mainly due to the lack of follow-up by scientific community and not due the growth in fraudulent research.

Explosive growth of uncited relations necessitates development of additional criteria for relation confidence besides the number of supporting references. We can suggest following approaches for such new confidence measure:

1. Adding epistemic discourse analysis to the edge annotation in the knowledge graph ^xii^
2. Adding article citation index to the relation confidence score.
3. Developing statistical score that measures overall alignment of a new reported relation with entire knowledge graph.
4. Additionally, cited research confidence scores can be further refined by distinguishing self-citation versus independent observations by other labs.

We can distinguish six credibility levels of published knowledge:

1. Highly cited scientific observations represent the golden set and the foundation of scientific knowledge used by textbooks.
2. Low cited scientific observations represent either emerging new knowledge or underfunded research areas. This data is also used in scientific books and review articles.
3. Uncited peer-reviewed scientific observations, which probably should include self-cited observations and data from high-throughput experiments.
4. Pre-print knowledge that did not pass peer review, but usually written by professional scientists and therefore have some degree of quality assurance.
5. Internet articles published by non-scientists, such as journalists, healthcare professionals, patients, and their families.
6. Due to generative AI, we anticipate significant growth of the content generated by large language models.

We argue that large language models that will be developed for the scientific community should be trained on different subsets of literature to allow for better sense of confidence of LLM output. LLMs trained using highly cited publications can be used to verify observations reported in low-confidence publications. Evaluation of low-confidence facts by high-confidence LLMs can assist human reviewers in the peer review process and set a path towards fully automated peer review.

## Conclusion

- Citation rate of facts reported in controlled access is no different from the citation rate of facts reported in open access.
- Half of all relations in the knowledge graph are supported only by one reference. These relations are equally distributed between CA and OA.
- Additional confidence scores are necessary to estimate the confidence of knowledge graph relations.

## Data Availability

All data produced in the present study are available upon reasonable request to the authors

## Abbreviations

EBKG: Elsevier Biology Knowledge Graph
OA: Open Access
CA: Controlled Access

## Author Contributions

All authors conceptualized, designed, and drafted the manuscript as well as provided critical review for important intellectual concepts and approved the final version to be published. All authors agree to be accountable for all aspects of the work.

## Acknowledgements

The authors would also like to acknowledge the contribution of Z Jelenje (Nascent Studio) who gave useful comments on the manuscript and guidance with typesetting and formatting.

Authors also thank Dr. Iryna Chelepis for her help in creating figures 10 and 11 as well as helpful review of the manuscript.

**Figure 11:**
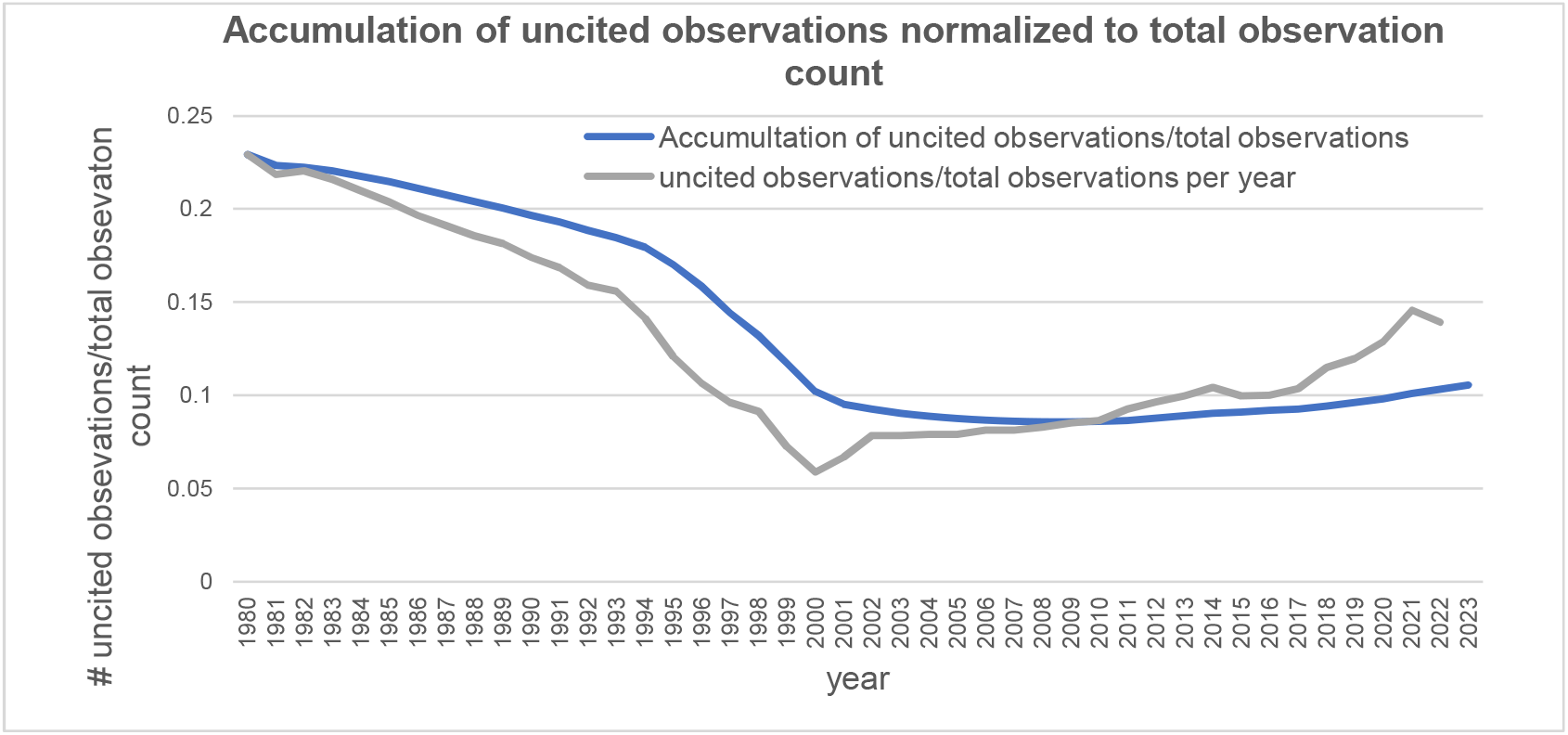
Yearly proportion of uncited observations. Accumulation graph shows the ratio of accumulated uncited observations relative to the total number of observations in EBKG every year.

**Figure 12:**
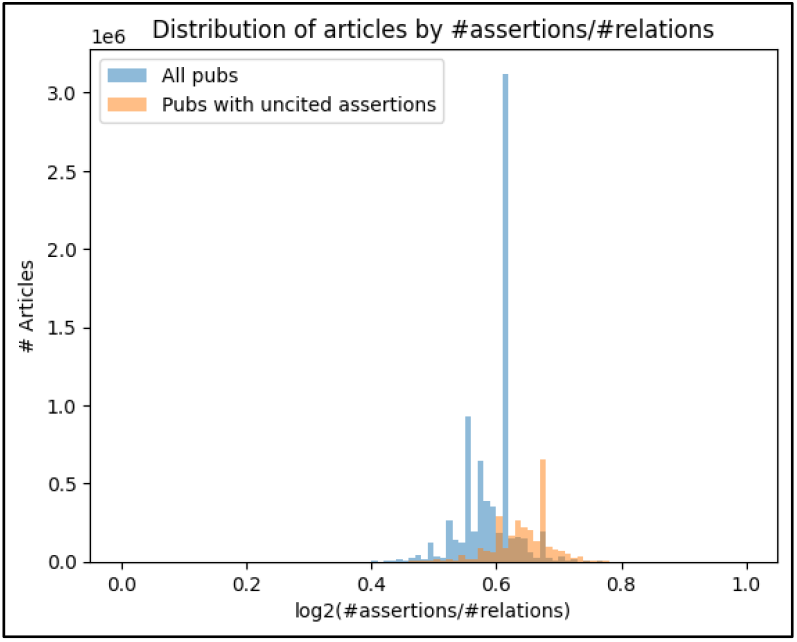
Comparing distributions of #assertions/#relations ratios among uncited observation articles versus all articles.

